# Perceptions of homogeneity reproduction in health sciences academia

**DOI:** 10.64898/2026.03.04.26347665

**Authors:** Rieke Buckup, Jamie Brian Smith, Gertraud Stadler, Pichit Buspavanich

## Abstract

Academic institutions privilege norms of continuous productivity and uninterrupted availability, creating conformity pressures that systematically disadvantage those who deviate from an implicit template of the ‘ideal academic’. This study explores how doctoral students and faculty in the health sciences perceive the reproduction of social homogeneity. Semi-structured interviews were conducted with nine participants at a German university hospital. Data were analysed using reflexive thematic analysis with extended idiographic engagement. Participants perceived homogeneity as reproduced through external exclusion, enacted by others through networks, normative expectations, or institutional arrangements, and self-exclusion, whereby individuals withdrew, reduced visibility, or reshaped identity in anticipation of exclusion (‘anticipatory compliance’). Across both processes, the tacit norm of the ideal academic organised access and belonging. Supportive supervision and visible role models were perceived as partial buffers but did not structurally alter underlying norms.

Interpreted through the social identity threat framework, these findings are consistent with a self-reinforcing cycle: structural homogeneity may generate identity-threatening environments that activate concealment and withdrawal, concentrating homogeneity further. These findings suggest that achieving substantive inclusion requires challenging the structural conditions that naturalise presence, mobility, and availability as measures of academic success.

## Introduction

Academic careers in the health sciences are widely organised around an image of the ‘ideal academic’: continuously productive, fully mobile, and unrestricted by care or bodily needs [1–5]. This practical norm shapes social homogeneity, expectations, and career advancement. The norm does not travel evenly across gender, race, class, disability, or family status, privileging those who can conform to an ideal and marginalising those who deviate [6,7]. The health sciences present a particularly instructive case: academic medicine layers clinical hierarchies onto academic ones, requires dual clinical and research paths that assume uninterrupted availability, and recruits from a narrow socioeconomic base [50]. In Germany, this dual-track structure is intensified by the Habilitation system, which requires a second major academic qualification beyond the doctorate, and by fixed-term contract legislation (Wissenschaftszeitvertragsgesetz) that limits pre-qualification employment to twelve years, compressing career timelines in ways that disproportionately penalise those with care responsibilities or non-linear biographies. Because the profession is public-facing, workforce homogeneity carries consequences for the diversity of perspectives brought to patient care and health research.

Research has documented persistent social homogeneity in senior roles, gendered and racialised barriers to career progression, and the mental-health costs of high-pressure academic cultures [2,5,8–11]. Accounts of a ‘leaky pipeline’ describe these patterned losses of participation and progression, while leaving open the question of how the ideal becomes enforceable in daily practice [12,13]. What remains less clear is the mechanism that links this norm to everyday experiences of homogeneity reproduction. In this article, we use conformity pressure as a sensitising concept for that mechanism [14–16]. Qualitative studies have documented these processes in specific domains, including gendered gatekeeping in professorial recruitment [11], racialised exclusion in the UK professoriate [8], and ableist assumptions in academic culture [6]. What this body of work has not yet offered is an account that traces how external exclusion and self-exclusion operate together as dual mechanisms of homogeneity reproduction, nor one situated in the specific conditions of German academic medicine. Drawing on interviews with doctoral students and faculty at a German university hospital, we examine how the reproduction of social homogeneity is perceived in everyday academic environments and how these perceptions relate to normative ideas about the ‘ideal academic’.

### The ideal academic: gendered organisations, exclusion, and the reproduction of social homogeneity

Universities are not gender-neutral spaces: They are built through concepts and routines that assume an abstract worker who can give total priority to work [1,2,19]. In higher education, this appears as the ideal academic: uninterrupted productivity, constant availability, and geographic mobility as enactments of commitment [3,20]. These ideals are materialised in evaluation rubrics and recruitment discourses of ‘excellence’, which calibrate inclusion and exclusion to particular temporalities and biographies [11]. Mobility expectations and international experience often operate as proxies for commitment and quality, despite uneven access to such opportunities [5]. Informal talk and tacit timelines then normalise these expectations in everyday judgements [7]. These cultures of evaluation, expectations of excellence, and networked homogeneity provide the conditions under which the ideal becomes practicable in academic careers.

The ideal is inflected by race, class, and disability. Actors in existing power networks favour those who resemble themselves, thus stabilising homogeneity over time [15,21–23], while gatekeeping in recruitment and networking reproduces dominant norms under the banner of excellence [11]. UK evidence documents racialised stratification at senior levels in Higher Education [8,24]. Classed assumptions about unpaid time, geographic mobility, and linguistic capital structure access to ‘fitting in’, and ableist norms treat certain bodies and neurotypes as excluded from academic time [6,7,25]. German studies further show the costs of care and mobility on publication and progression. In Germany, women publish 20% less than men [26].

### From ideals to practice: conformity pressure as mechanism

Conformity pressure refers to the diffuse, routine pressures through which certain behaviours, bodies, and biographies come to signify academic commitment [14,25]. We use conformity pressure as a sensitising concept in Blumer’s sense: it directed our analytical attention to how norms are enforced in daily practice without predetermining the forms that enforcement takes [14]. Networks and institutional hierarchies that heighten scrutiny for those who do not conform to prevailing templates intensify these pressures [11,15].

Social identity threat offers a complementary account of why conformity pressure is experienced as threatening by those who deviate from the dominant template. Developed within the social identity tradition, the construct describes the experience of appraising a situation as potentially harmful to the value or enactment of a social identity one holds [29,30]. Branscombe and colleagues [29] distinguish four forms: categorisation (being reduced to group membership), distinctiveness (erasure of group boundaries), value (devaluation of one’s group), and acceptance (exclusion from a group one considers oneself part of). In academic environments organised around the ideal academic, those whose social identities mark them as non-normative may experience acceptance threat and value threat simultaneously: their belonging is uncertain and their group membership is treated as incompatible with academic commitment. The identity management strategies that follow (vigilance, concealment, compensatory overperformance, withdrawal) overlap with the minority-stress processes documented among minority scholars [16] and specify the situational mechanism linking conformity pressure to exclusion practices. We return to this framework and to Schmader and Sedikides’s [61] State Authenticity as Fit to Environment (SAFE) model in interpreting our findings.

### Consequences, coping, and moderating conditions

Under sustained conformity pressure in institutions, people mobilise different repertoires, including exit/avoidance, conformity and compensation [31–33]. Exit or avoidance reduces exposure to judgement (e.g., avoiding particular labs, opting for lower-visibility roles) but narrows opportunity structures [32,34]. Attempted conformity mutes differences to reduce visibility (e.g., silence around care, disability, or migration histories), often at the cost of authenticity and health [6,16]. Compensation through overperformance buys credibility while elevating workload, perfectionism, and strain [33,35]. Empirical syntheses of doctoral mental health corroborate these costs, documenting elevated anxiety, depression, and suicidality relative to general populations, with role demands, uncertainty, and stigma-related presenteeism as key drivers [9,35,36].

Supervision can widen the terms of belonging by making expectations explicit, calibrating criteria to developmental stage, and broadening what counts as contribution [17,18,33]. Visible role models who diverge from the dominant template can provide local support; however, they rarely re-write organisational logics on their own.

### Research questions

How do faculty and doctoral students perceive the reproduction of social homogeneity in academic environments?

How are academic norms articulated and made visible through these perceptions?

## Materials and methods

### Research design

This study explores experiences of doctoral students and faculty members at a German university medical school, as part of a broader mixed-methods project investigating diversity and opportunity in academia. We adopted reflexive thematic analysis within an interpretivist frame to develop themes about normative orders, complemented by extended idiographic engagement with contrasting cases [38]. The study adhered to COREQ guidelines (S5 Checklist).

### Analytical approach

Our primary method was reflexive thematic analysis (RTA) [39] to map shared patterns across interviews. We moved from familiarisation and generative coding to theme development, using reflexive memos and analytic conversations to surface difference and check assumptions. Themes capture how the ‘ideal academic’ is named, how perceived social homogeneity makes that norm visible, and how everyday practices and arrangements enforce it. This stage provides the cross-case architecture of the analysis.

To add idiographic depth, we undertook extended line-by-line engagement with two analytically rich, contrasting interviews, drawing on the idiographic sensibility of interpretative phenomenological analysis [38] to trace personal meaning-making around discrimination, coping, and boundary-setting. This idiographic work deepens rather than duplicates the thematic account: it elaborates how the processes identified via RTA are lived, rather than generalising from cases. Cross-case comparison within this idiographic stage was limited to clarifying how the identified mechanisms operate, rather than seeking convergence across participants.

### Reflexivity

Interviews were conducted by a female, bilingual (German/English) doctoral researcher (MSc) with a background in social science and medicine, formally trained in qualitative methods. We acknowledge that the interviewer’s positionality, being white, cisgender, and an early-career academic with explicit diversity advocacy commitments, influenced the research process. This created a complex insider-outsider dynamic: shared institutional affiliations and experiences with participants potentially facilitated rapport while risking assumptions of shared understanding, affecting both participants’ narrative framing and researcher interpretation [40].

To mitigate biases, we employed reflexive journaling, peer debriefings, and analytic memoing throughout data collection and interpretation, to support analytic transparency and critical interrogation of positionality’s influence. No pre-existing relationships existed with participants recruited through snowball sampling. While participants knew the researcher’s affiliation with the Advance Academia project, potentially influencing their accounts, we view positional reflexivity not as limitation but as necessary and generative for researching sensitive topics like discrimination and inclusion in academia [40].

### Participant selection

Initially, we targeted a stratified sample representing different career stages and diversity characteristics using a recruitment matrix based on publicly available diversity data [41] (see S4 Table). After limited success with email recruitment, we shifted to opportunistic snowball sampling. Initial participants from researchers’ professional networks recommended others meeting the inclusion criteria: current doctoral students or faculty members affiliated with the medical school. Snowball sampling risks homogeneity, as participants typically recommend individuals from similar circles, potentially underrepresenting certain identity groups. Of 20 individuals approached, two explicitly declined and nine did not respond after one follow-up; the remaining nine participated.

Sample adequacy was judged using Malterud and colleagues’ [62] concept of information power. Our narrow aim (perceptions of homogeneity reproduction in one institutional context), the specificity of the sample (all participants held direct experience of the phenomenon), the use of an established theoretical lens (social identity threat), the depth of dialogue achieved (45–75 minute interviews yielding rich, reflexive accounts), and the detail of our analytic strategy together supported adequate information power for the research questions. The final sample included nine participants: three doctoral students (all women) and six faculty members (three women, three men), as summarised in Table 1. Participants self-disclosed a range of diversity-relevant characteristics during interviews, including sexual orientation, neurodiversity, care responsibilities, class background, and migration history. To protect anonymity in this single-institution sample, these characteristics are not linked to individual participants in Table 1 but are reflected in the quotations presented in the findings.

**Table 1.**
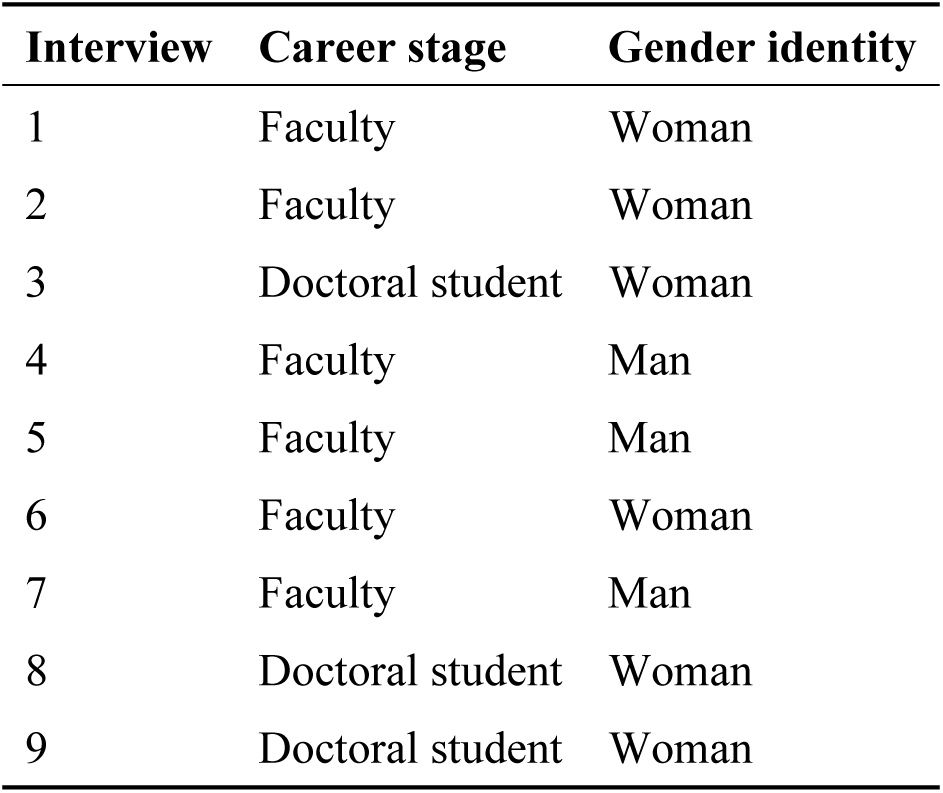
Participant overview.

| Interview | Career stage | Gender identity |
| --- | --- | --- |
| 1 | Faculty | Woman |
| 2 | Faculty | Woman |
| 3 | Doctoral student | Woman |
| 4 | Faculty | Man |
| 5 | Faculty | Man |
| 6 | Faculty | Woman |
| 7 | Faculty | Man |
| 8 | Doctoral student | Woman |
| 9 | Doctoral student | Woman |

### Setting and data collection

Individual, semi-structured interviews were conducted either in-person at the university hospital or via video conferencing, depending on participant preference and availability. No individuals other than the interviewer and participant were present during the interviews. All interviews (45–75 minutes) were conducted in German, securely audio-recorded, and transcribed verbatim by the research team. The interviewer made brief field notes after each interview to record contextual observations and initial analytic reflections.

The interview guide (S1 Appendix) covered topics related to the academic environment. The guide was piloted within the research team and refined based on their feedback. Participants were recruited and interviewed between September 2024 and December 2024. No repeat interviews were carried out, and transcripts were not returned to participants for comment.

Participants were not invited to provide feedback on the findings, consistent with our reflexive thematic analysis approach, which positions the researcher as an active interpreter of meaning rather than a neutral reporter of participant views [39]. Transcripts were translated into English by the bilingual interviewer. A second bilingual team member independently back-translated selected passages to verify accuracy and preservation of meaning; discrepancies were resolved through discussion. Quotations presented in this article retain the translated phrasing and are marked with the interview number and participant role.

## Data analysis

To address both overarching patterns and the specifics of individual meaning-making, we conducted reflexive thematic analysis following Braun and Clarke [42], generating themes inductively while remaining attentive to the context and complexity of participants’ accounts. Two researchers coded transcripts independently, then engaged in iterative analytic dialogue to refine interpretations using MAXQDA software [43,44]. This process was not aimed at consensus but at deepening and challenging each researcher’s reading through collaborative discussion, consistent with the reflexive approach [39]. We then pursued extended idiographic engagement with two contrasting cases, drawing on the idiographic sensibility of IPA [38], to trace how the thematic patterns were lived and made sense of by individual participants. The two cases selected for extended engagement were Interview 4 (a faculty member whose account articulated the normative force of the ideal academic from an enacting perspective) and Interview 6 (a faculty member whose account traced the cumulative personal costs of managing multiple non-normative identities). Interview 4’s enacting perspective is woven throughout the thematic analysis because it recurs across multiple external exclusion themes; Interview 6 receives a dedicated idiographic section because the convergence of multiple non-normative identity dimensions required sustained biographical attention. An example of a coding tree is available in S3 Appendix.

## Ethical considerations

Ethical approval for this study was obtained from the Ethics Committee of Charite – Universitatsmedizin Berlin (reference number: EA1_034_22). All participants provided written informed consent prior to their interview (see S2 Appendix). Potentially identifying information has been removed from quotes. The study is sponsored by the Berlin University Alliance.

## Data availability

The qualitative interview data underlying this study cannot be made publicly available because they contain potentially identifying information about participants employed at a single institution. Sharing these data would violate the terms of informed consent and the conditions of ethics approval (EA1_034_22). De-identified excerpts relevant to the findings are presented within the article. Requests for access to additional anonymised data excerpts may be directed to the Ethics Committee of Charite – Universitatsmedizin Berlin.

## Results and discussion

Across interviews, exclusion emerged as the organising construct through which participants made sense of homogeneity reproduction. The norm of the ideal academic is both institutionalised and internalised, organising exclusion at individual and structural levels. We treat exclusion as a second-order analytic construct, drawing on both participant accounts and, where relevant, statistical evidence to contextualise their perceptions. This deliberate layering of statistical context within an interpretivist frame treats such evidence as contextualisation, not corroboration, situating participants’ meaning-making within the broader structural conditions they inhabit.

We distinguish external exclusion from self-exclusion. External exclusion refers to gatekeeping, conscious or otherwise, enacted by individuals, groups, or institutions: not only explicit denial of access but also structural and discursive processes that disadvantage those who do not match normative expectations [11]. Self-exclusion refers to processes in which individuals, under conformity pressure, withdraw, reduce visibility, or reshape identity in anticipation of exclusion. We term this pre-emptive reshaping ‘anticipatory compliance’: it encompasses covering [57] and passing [31] but extends to the reshaping of behaviour, career plans, and self-presentation before any specific threat materialises. The concept shares ground with Foucauldian self-surveillance, but differs in that our participants described conscious, strategic reshaping of identity rather than the pre-reflective self-policing that disciplinary accounts emphasise. Where Meyer’s [16] minority stress model identifies expectations of rejection among sexual minorities, anticipatory compliance extends this mechanism across gender, class, disability, neurodiversity, and care status. Unlike accounts that treat self-exclusion as full opt-out from career paths [45], we include partial practices that exclude aspects of identity rather than the person from academia altogether. The two processes are intertwined, with those affected holding varying degrees of agency.

## External exclusion

Participants perceived external exclusion as operating across interpersonal and institutional levels: in access to leadership positions, in scholarly networks, and in organisational and physical arrangements. Interviewees noted:

> For example, in committees where there are simply a lot of men who then vote for other men, or where only men are nominated in the first place and then people say, well, no women nominated themselves, and you think, okay, but maybe no women felt inspired to do so (Interview 3, student).

> It is difficult to appoint women to professorships because, in my experience, the fact that women are to be given preference if they have the same qualifications means that they are actually disadvantaged. In other words, they are not even invited in the first place. Because as soon as they are invited, they can be given preference if they have the same qualifications (Interview 7, faculty).

The persistent male dominance accords with literature on homophily and homosocial reproduction, whereby decision-makers prioritise those most similar to themselves [47].

Statistical evidence on the broader composition of medical academia suggests that restricted access to leadership positions is a structural pattern. For example, 75.8% of senior researchers in German medical academia are male [48], while women occupied only 10% of leading university hospital positions as of 2016 [49]. Socioeconomically, 57.9% of doctors and 73.8% of medical students come from the upper wealth quintile, with lower quintiles largely underrepresented [50]. Turkish and Polish communities, among the largest migrant groups in Germany, are underrepresented among clinicians, consistent with international evidence that medicine disproportionately recruits from higher-income, majority-ethnic families [51]. The university hospital context compounds these patterns: academic medicine requires managing parallel clinical and research hierarchies, in which progression depends on scholarly output and on clinical credentialing pathways that assume full-time, uninterrupted availability. Clinical service and academic production together intensify conformity pressure, narrowing the template further than in non-clinical disciplines. The costs of non-conformity are acute: reduced clinical hours, interrupted training, and lost access to patient-facing roles that generate research output.

External exclusion extended to the structuring of academic networks. One faculty member described networks as “the closed ranks of colleagues…the networks of old men” (Interview 4, faculty). Another faculty member noted: “When I look at my networks, it’s fair to say […] they’re almost all native German speakers with white skin colour” (Interview 5, faculty). Academic networks are thus experienced as socially homogeneous and exclusive in their access conditions. In the Weberian sense, this perceived dynamic constitutes ‘social closure’: an academic elite restricting access to outsiders and reproducing privilege through networked homogeneity [21,22,46].

External exclusion also operated through normative expectations that implicitly defined who could belong. One such mechanism was the ability to ‘speak’ academic language. As one faculty member noted:

> I think it really makes a difference, just because it’s a completely different language and a different self-understanding that some people already bring along from the beginning, because they got that from home (Interview 1, faculty).

Such expectations ‘marked’ those who deviated as incompatible [8] and were routinely voiced to newcomers, particularly doctoral students, who are presumed to be productive, constantly available, mobile, and free from care or other personal commitments. As one faculty member noted:

I am occasionally annoyed, I have to say that. When people suddenly realise that they have to have two children […] just before their third dissertation is due, and then their whole career is put on hold again […] So sometimes I get the impression that young women don’t plan enough. But maybe it’s presumptuous from my point of view to demand it like that…So quite simply young women in love who have somehow just got married. Suddenly they think now I want a child, and then their career is suddenly gone. That kind of thing too. These kinds of irrationalities also exist (Interview 4, faculty).

Parenthood is constructed as an ‘irrational’ deviation, implying that becoming a parent happens suddenly or impulsively. The frustration expressed toward female doctoral students whose caring responsibilities are seen as incompatible with expectations of constant performance frames them as failing to meet norms of continuous productivity [3]. Labelling it ‘irrational’ works to delegitimise and suggest a lack of forethought, thereby reproducing long-standing dualisms that position women as irrational and men as rational [52]. The faculty member does not problematise this exclusion normatively but, instead, treats institutional regulations as given, calling on women to adapt to them. The faculty member recognises the very same power structures described by other participants (i.e. being excluded when becoming a parent) but frames them not as unjust structures but as rules to accommodate to. We interpret this framing as a mechanism of external exclusion, whereby women who do not conform to academic norms are not explicitly excluded but rather marked as unreliable, thereby legitimising their exclusion.

External exclusion further manifested through institutional arrangements.

Interviewees pointed to the absence of accessible facilities for people with physical disabilities as a mechanism reproducing homogeneity. As two faculty members articulated:

> I think it is extremely difficult for people with physical disabilities […]. Congresses are more difficult to access or not at all. Depending on the type of physical ability, certain forms of communication are excluded, and it’s just not easily possible (Interview 5, faculty).

> The lift here has been broken for months now. You wouldn’t be able to get in here as a disabled employee or as a disabled wheelchair user (Interview 4, faculty).

Ableist infrastructures rendered full participation practically impossible. Such neglect constitutes normalised ableism [6]. External studies report comparable patterns and link non-accommodating academic environments for people with physical disabilities to psychological harm, including anxiety and depression [27,28].

Faculty and doctoral students perceived current educational systems, including curricula and regulatory frameworks, as inadequately accommodating neurodiverse individuals. Neurotypical norms meant that only those who could bear stress and suppress their needs remained competitive in the academic system. As faculty members noted:

> We increasingly have […] clearly neurodiverse students, which the system is not designed for at the moment […] And there’s a kind of mainstream stamp on it and I can sometimes get it to expand a bit with the resources I have available, but it’s extremely difficult (Interview 5, faculty).

> I’ve done a lot of things, and I pushed myself past myself boundaries […] in terms of that stress and accumulated stress […] ignoring and denying and pushing away needs […] for healing, uh, neglecting needs, neglecting relationships, all sorts of things that have harmed me (Interview 6, faculty).

External research supports these perceptions, showing that medical academia is largely structured around the ‘survival of the most abled’ [53]. By prioritising standardised approaches to learning and progression, academic institutions create a fundamental mismatch between neurodiverse needs and existing educational frameworks [53,54]. The second quote, from an interviewee identifying as neurodiverse, illustrates the personal strains of working within such an environment. Scholarship on neurodiversity in higher education documents associations between neurodiverse status and heightened risks of stress, depression, anxiety, and burnout [54].

Lastly, interviewees perceived that people with caregiving responsibilities face institutional exclusion:

> If I have childcare commitments or a person in need of care at home and actually need to go to a scientific conference? Nobody has an answer to that. So is there any kind of offer? I think relatively little (Interview 5, faculty).

> It’s quite clear that it’s a career stopper to a certain extent or a stumbling (Interview 2, faculty).

One cis-male-identified faculty member confirmed how institutional structures privilege uninterrupted availability: “My wife has taken parental leave. I didn’t (…) but it helped me a lot that I didn’t take parental leave, so I continued to work” (Interview 7, faculty). External exclusion thus operates through the absence of institutional arrangements as much as through active gatekeeping. Acker [1] calls this the ‘gendered organisation’, in which care responsibilities are rendered ‘irrationalities’ and academic careers follow hegemonically masculine paths [23], whereby precarious contracts, mobility demands, and intensive work culture systematically disadvantage caregivers, typically women [55]. The forms of external exclusion described here (restricted access to networks, normative marking of non-conformity, and infrastructural neglect) correspond to acceptance threat and value threat [29], creating the conditions under which self-exclusion becomes intelligible.

### Self-exclusion

Exclusion also operated from within. Under persistent conformity pressure, some interviewees described withdrawing from opportunities or limiting their visibility. Returning to the committee example, the interviewee noted that “no women nominated themselves, and you think, okay, but maybe no women felt inspired to do so” (Interview 3, student): a form of self-exclusion that involves agency (the decision not to apply) while being shaped by structural conditions (repeated messages of not belonging) that encourage withdrawal. In male-dominated settings, the mere absence of representation may deter potential applications: women may choose not to put themselves forward not for lack of ambition but because the conditions feel unwelcoming. External exclusion and self-exclusion are mutually reinforcing: structural homogeneity produces the conditions under which withdrawal becomes intelligible, and withdrawal in turn deepens the homogeneity that prompted it.

Self-exclusion also manifested as anticipatory compliance. A faculty member noted: “I live alone and don’t have children. I think if children had been involved at the time, I wouldn’t have taken this path” (Interview 1, faculty). This anticipation aligns with the ‘leaky pipeline’ [12]: whilst women form the majority of early-stage healthcare professionals in Germany [56], men dominate leadership roles. One student observed: “At assistant level, there are still more women and at some point there will be more men…especially in positions of responsibility” (Interview 8, student). The pipeline metaphor, however, locates the problem in individual departure rather than in the structures that produce departure. Another doctoral student captured the gap between institutional rhetoric and lived reality: “we’re always told that it’s all possible and so on, but in fact, it’s often not possible” (Interview 9, student).

These partial practices took several forms. Covering was central [57]: downplaying social identity markers to avoid being labelled ‘too different’. As one student explained:

> I think I have the feeling that the survival strategy of many marginalised people […] is to reduce the projection surface and that the best way to get through is to offer little projection surface (Interview 8, student).

Similarly, one faculty member described the hesitation to disclose queerness:

> A kind of typical life situation is expected (…) I still think about how openly I deal with being gay in work contexts, being queer…I would be very reluctant to say I was with my partner (…) while everyone else is saying I was there with my girlfriend. I (don’t) have any contact and no (…) lubricant for social contacts in the academic field (Interview 5, faculty).

Conformity pressures to align with heteronormative expectations prompt anticipatory withdrawal, which in turn reproduces homogeneity. In highly homogenous networks, interviewees experienced how tacit expectations about the ‘acceptable’ relationship subtly reward those who bring a heterosexual partner and nudge others to conform or cover.

Where covering involved suppressing non-normative aspects of identity, conformity pressure also operated through the proactive adoption of dominant norms. A faculty member noted:

> [Women], who have learnt, for example, to be able to practise the networks like men, who put their own family on the back burner, if they have one at all, i.e. who then become career women, who then also manage to get into these higher positions. The Charite also used to have a female dean. So, it’s not as if that hasn’t happened. Yes, and there are women in management positions here too, but I think they pay a very high price (Interview 4, faculty).

Women who want to succeed academically, this faculty member suggested, must conform to behaviours typically associated with cis-male behaviour and adopt hegemonically masculine norms [58], to, in effect, ‘become male’. Conformity here operates as self-exclusion: proactively reshaping or abandoning parts of identity.

A further dimension of self-exclusion was compensatory overperformance. Where participants could not conform to the ideal academic on dimensions such as gender, sexuality, neurodiversity, or care responsibilities, they described compensating by exceeding expectations on the dimensions they could fulfil. Overperformance functions as anticipatory compliance: working to be seen as acceptable by subordinating other facets of identity to output. As faculty members noted:

> If I am weird then I have to be magical, but I can guarantee delivery (…) no matter what (…) I can solve impossible problems (Interview 6, faculty).

> I did feel even from like entering academia each year that I needed to be perfect to be able to exist as a queer person (Interview 6, faculty).

> There are women in leadership positions, but I think they pay a very high price (…) they always have to go one better, always have to go the extra mile (Interview 4, faculty).

Overcompensation may ultimately reinscribe academic ideals of constant output, reinforcing the very standard it seeks to overcome [3]. Anticipatory compliance, alongside covering, further contributes to minority stress through the ongoing demand to justify or compensate for one’s identity [16]. That participants described these strategies as routine rather than exceptional suggests that identity threat operates as a structural feature of environments organised around the ideal academic.

The account of one faculty member (Interview 6) illustrates how these processes converge in a single biography. This participant, who identified as both neurodiverse and queer, described accumulated harm from suppressing needs to meet academic demands, ‘ignoring and denying and pushing away needs […] for healing, neglecting needs, neglecting relationships, all sorts of things that have harmed me’, while simultaneously performing exceptional competence to secure belonging: ‘If I am weird then I have to be magical.’ For this participant, the ideal academic was not an abstract norm but a daily negotiation in which multiple non-normative identities compounded the pressure to compensate. The felt need to ‘be perfect to be able to exist as a queer person’ reveals how identity threat operates across dimensions simultaneously: queerness, neurodivergence, and the demand for continuous output intersected to produce a mode of academic survival experienced as both necessary and personally costly. This extended engagement with a single case illustrates how the thematic patterns identified via reflexive thematic analysis are not experienced as discrete processes but as dimensions of a single lived reality.

### Moderating conditions: Role models and supervision

Interviewees emphasised the value of seeing academics who deviate from hegemonic norms in visible posts. For some, the presence of women combining academic work and family expanded what felt feasible. One participant remarked: “Seeing my supervisor, who is both a dedicated mother and a brilliant academic, helped me believe that it’s possible to balance both” (Interview 8, student). Yet, as Spaans et al. [59] suggest, minorities tend to not have demographically similar role models. One faculty member stated: “The higher you go, the fewer women you see in leadership” (Interview 1, faculty), indicating that role models might only rarely occur in positions of power. Although interviewees perceived role models as widening their imagery of belonging, in contexts of persistent homogeneity their effects are uneven and contingent. This can obscure the structural conditions underlying academic norms. From a social identity threat perspective [29], visible role models may reduce acceptance threat locally by signalling that non-normative identities can belong in academic spaces, yet without structural change, such signals remain individual-level exceptions.

Interviewees also perceived good supervision as important for reducing feelings of isolation and mitigating the perceived demands occurring with academic norms. One doctoral student emphasised:

> I can always see if there are any summer schools or similar on methodology or content, and then ask at the institute whether there are funds available or through the project. And I am actually quite well supported in that regard (Interview 3, student).

Good supervision here extended beyond the interpersonal to alleviate structural barriers such as financial access to further education. Literature suggests that supervision is central to the wellbeing of doctoral students and early-career academics, particularly for those who simultaneously experience minority stress [16,37]. Yet the quality of supervision depends strongly on individual factors and can hardly be guaranteed on an institutional basis [60]. While some interviewees described good supervision as essential, it remains contingent and precarious. A doctoral student captured this contingency: “(the previous institute head) has just left, and now a new one is coming in. We’ll have to see how things continue from here” (Interview 3, student). Good supervision is thus not institutionally guaranteed but depends on the commitment of individuals, raising the question of whether it alleviates structural problems or, at times, obscures them. As with role models, supervision may buffer individual experiences of conformity pressure without altering the norms from which exclusion arises.

## Limitations

The findings of this study should be interpreted in consideration of its limitations.

First, the single-institution sample may limit the transferability of the findings to other contexts. Second, snowball sampling likely produced a more homogeneous sample, as participants tend to refer to others within similar networks, potentially excluding marginalised or isolated individuals. The 55% non-response rate (eleven of twenty individuals approached) warrants reflexive consideration: in a study about exclusion and silencing, those who did not participate may include individuals who experience the most acute marginalisation, and their absence may narrow the range of experiences captured. Third, our sample does not include participants who identified as racially or ethnically minoritised, nor individuals with physical disabilities, limiting what we can say about these dimensions of exclusion. Fourth, faculty outnumber students six to three, which may privilege the enacting perspective over the receiving perspective in our account. Fifth, we did not explicitly search for specific diversity domains, relying on self-report, which may have limited our analytical scope. Finally, researchers’ experiences and preconceptions, including the insider-outsider positionality described above, may have influenced participants’ disclosures and our thematic emphases despite reflexivity efforts.

## Conclusions

Our findings identify exclusion, external and self-directed, as the mechanism through which social homogeneity is reproduced in academic environments. Role models and supervision buffered individual experiences but left underlying norms intact.

Read through the social identity threat framework [29,30], our findings are consistent with a self-reinforcing cycle. External exclusion contributes to environments in which non-normative identities are devalued, which may generate acceptance threat and value threat that activate the identity management strategies our participants described. When these strategies prove insufficient, withdrawal further concentrates homogeneity; the cycle may begin again with a yet more homogeneous environment that intensifies identity threat for those who remain.

Schmader and Sedikides’s [61] model of State Authenticity as Fit to Environment (SAFE) clarifies the mechanism linking identity threat to withdrawal. The model proposes that individuals experience authenticity when three conditions of person-environment fit are met: self-concept fit (alignment between self-perception and how one is perceived), goal fit (the environment supports one’s aims), and social fit (a sense of belonging among peers). Our participants’ accounts correspond to these dimensions: faculty and doctoral students who perceived themselves as marked by gender, sexuality, care responsibilities, disability, or class background described environments in which they could not be fully themselves, could not pursue academic goals on equal terms, and felt excluded from the networks through which belonging is enacted. The anticipatory compliance we observed can be understood as attempts to manufacture provisional fit in environments that do not afford authenticity to non-normative identities. Self-exclusion, in this framing, is not a failure of individual ambition but a rational response to sustained inauthenticity; people withdraw from settings in which they cannot be who they are. The SAFE model assumes a stable authentic self against which fit can be measured; for participants managing multiple non-normative identities simultaneously, as Interview 6 illustrates, the self may be plural and situationally constituted, complicating any single measure of fit.

We identify four implications for policy and practice. First, decoupling presence, mobility, and temporal intensity from assessments of success by recognising non-linear careers to accommodate care and other responsibilities. Second, investing in accessible infrastructures, including physical (e.g., lifts, conference access), organisational (childcare, flexible timetables), and procedural (transparent workload models, explicit criteria early in training). Third, discussing underlying norms in supervisory practice, clarifying expectations and legitimate boundaries of availability. Finally, recognising neurodiversity and disability not as exceptions to be endured but as dimensions around which learning environments are designed. Our single-site design situates claims rather than generalises them. Future work should examine whether reducing conformity pressures disrupts the recursive reproduction of social homogeneity, including decreasing minority stress. The central implication is straightforward: changing who thrives in academia requires changing what counts as academic thriving.

## Supporting information

**S1 Appendix. Interview guide.** Semi-structured interview guide used for data collection.

**S2 Appendix. Participant consent form.** Informed consent form provided to all participants prior to interview.

**S3 Appendix. Coding tree.** Example of the coding tree developed during reflexive thematic analysis.

**S4 Table. Recruitment matrix.** Recruitment matrix showing targeted and achieved sample characteristics.

**S5 Checklist. COREQ checklist.** Completed Consolidated Criteria for Reporting Qualitative Research (COREQ) 32-item checklist.

## Data Availability

Please email the authors for discussions about data access

